# Predictors of COVID-19 vaccine uptake: An online longitudinal study of US Veterans and non-Veterans

**DOI:** 10.1101/2022.04.19.22273818

**Authors:** Alistair Thorpe, Angela Fagerlin, Frank A. Drews, Holly Shoemaker, Federica S. Brecha, Laura D. Scherer

## Abstract

**Background:** To effectively promote vaccine uptake, it is important to understand which people are most and least inclined to be vaccinated and why.

**Purpose:** To identify predictors of COVID-19 vaccine uptake and reasons for non-vaccination.

**Design:** A longitudinal English-language survey study.

**Setting:** Online in December-2020, January-2021, and March-2021. Participants. 930 US respondents (63% Veterans).

**Measurements:** Surveys included questions about respondents’ behaviors, well-being, healthcare experiences, and attitudes regarding the pandemic.

**Results:** The proportion of respondents who received ≥1-dose of a COVID-19 vaccine increased from 18% in January to 67% in March. Older age predicted vaccine uptake in January (OR=2.02[95%CI=1.14–3.78], *p*<.001) and March (10.92[6.76–18.05], *p*<.001). In January, additional predictors of vaccine uptake were higher numeracy (1.48[1.20–1.86], *p*<.001), COVID-19 risk perceptions (1.35[1.03–1.78], *p*=.029), and believing it is important that adults get the COVID-19 vaccine (1.66[1.05–2.66], *p*=.033). In March, additional predictors of vaccine uptake were believing it is important that adults get the COVID-19 vaccine (1.63[1.15–2.34], *p*=.006), previous (January) COVID-19 vaccine intentions (1.37[1.10–1.72], *p*=.006), and belief in science (0.84[0.72–0.99], *p*=.041). Concerns about side effects and the vaccine development process were the most common reasons for non-vaccination. Unvaccinated respondents with no interest in getting a COVID-19 vaccine were younger (0.27[0.09–0.77], *p*=.016), held negative views about COVID-19 vaccines for adults (0.15[0.08–0.26], *p*<.001), had lower trust in healthcare (0.59[0.36–0.95], *p*=.032), and preferred to watch and wait in clinically ambiguous medical situations (0.66[0.48–0.89], *p*=.007).

**Limitations:** Reliance on the accuracy and consistency of self-reported data.

**Conclusion:** These findings offer important insights regarding key predictors of vaccine uptake during the early stages of the COVID-19 vaccine rollout in the US, which can help guide health communications and public outreach. Evidence that attitudes and intentions towards COVID-19 vaccines are important predictors of uptake provides validation for studies which have used these measures and reinforces the need to develop effective strategies for addressing concerns about vaccine safety and development which continue to be at the forefront of vaccine hesitancy.

**Registration:** The pre-registration document associated with this manuscript is available at: https://aspredicted.org/MKS_HRZ.

## Introduction

COVID-19 continues to pose a significant threat to public health. Widespread uptake of the multiple vaccines authorized by the U.S. Food and Drug Administration for use against COVID-19 represents the safest and most effective strategy for limiting the impact of the disease.^1^ However, public hesitancy and refusal to get vaccinated remains a major challenge to realizing the full preventative health benefits of the authorized COVID-19 vaccines.^2^

In order to effectively promote vaccine uptake, it is important to first understand which people are most and least inclined to be vaccinated and why. Over the course of the pandemic, research identifying important demographic (e.g., age, race, ethnicity, and education)^2–5^ and psychological factors (e.g., COVID-19 risk perceptions,^3,4,6,7^ belief in conspiracy theories,^8^ political affiliation,^3,6,7,9^ exposure to misinformation,^10^ and trust in scientists,^6,11^ and the government^11,12^) associated with public attitudes and intentions towards COVID-19 vaccines has accumulated at a rapid rate. This research has been of great value to policy makers and health communicators aiming to develop strategies and interventions to address concerns about COVID-19 vaccines and promote vaccine uptake.

However, while attitudes and intentions towards vaccination are often useful predictors of behavior^13–15^ they do not always translate into actual vaccine uptake.^16–20^ For example, it is well documented that many people who intend to receive an influenza vaccine ultimately do not go on to receive one.^16,18^ Longitudinal data is therefore needed to identify attitudinal and sociodemographic factors that predict future vaccine uptake. The aim of the present study is to identify factors that predicted uptake of COVID-19 vaccination when vaccines first became available in January and March-2021, and to report the reasons given for not getting vaccinated by those who had not and did not intend to, following the rollout of the COVID-19 vaccines in the US in December-2020.

We expected that older age, living in a state with a greater proportion of people vaccinated, Veteran status, having a greater number of pre-existing health conditions, higher health literacy, higher numeracy,^a^ and being non-Hispanic white, would be associated with having received at least one dose of a COVID-19 vaccine in both January and March-2021. Based on existing research on psychological factors associated with COVID-19 vaccine attitudes and intentions, we also expected that greater worry about COVID-19, greater COVID-19 risk perceptions, greater confidence in vaccines, greater intentions to get a COVID-19 vaccine, greater trust in health care, greater belief in science, less belief in conspiracies, more liberal political views, and medical maximizing would be associated with COVID-19 vaccine uptake.

## Method

### Recruitment and respondents

Respondents were recruited and compensated by Qualtrics Online Panels for three surveys as part of a longitudinal study conducted in December 2-27, 2020 (*n*_Veteran_=1060; *n*_nonVeteran_=1025), January 21-February 6, 2021 (*n*_Veteran_=746; *n*_nonVeteran_=511), and March 8-23, 2021 (*n*_Veteran_=688; *n*_nonVeteran_=387). The surveys were in presented in English and administered online. This study was deemed exempt by the University of Utah and the Salt Lake City VA IRBs and follows the reporting guidelines of the American Association for Public Opinion Research.

A total of 930 respondents completed all three surveys and were included in the analyses. Information on the 1,155 respondents who did not complete all three surveys is available at: https://rpubs.com/AThorpe/CV19VA_Dropouts. The completion rate was 44% overall, 55% for Veterans, and 33% for non-Veterans. Respondents in our sample were generally older (median age ranged between 55 and 74 years old), male (735 (79%)), non-Hispanic White (720 (77%)), US Veterans (584 (63%)), and with a median household income between $50,000-$99,999. Over half of respondents (440 (64%)) reported having a pre-existing condition that made them more vulnerable to COVID-19; 186 respondents (27%) indicated that they did not have such a pre-existing condition and 67 respondents (10%) were not sure. In January, 165 respondents (18%) reported having received a COVID-19 vaccine; 160 (97%) of those were first doses and only 5 (3%) had received both doses. The number of respondents reported having been vaccinated increased to 620 (67%) in March with 206 (33%) first doses and 414 (67%) both doses. Full demographics are shown in Table 1.

**Table 1.** Respondent demographics overall and according to Veteran status.

|  | Overall<br>(n=930) | Veteran<br>(n=584) | Non-Veteran<br>(n=346) |
| --- | --- | --- | --- |
| Age — yr |  |  |  |
| 18 to 34 | 37 (4%) | 0 (0%) | 37 (11%) |
| 35 to 54 | 87 (9%) | 16 (3%) | 71 (21%) |
| 55 to 74 | 591 (64%) | 390 (67%) | 201 (58%) |
| 75 or older | 213 (23%) | 176 (30%) | 37 (10%) |
| Did not respond | 2 (<1%) | 2 (<1%) | 0 (0%) |
| Gender |  |  |  |
| Female | 193 (21%) | 38 (7%) | 155 (45%) |
| Male | 735 (79%) | 545 (93%) | 190 (55%) |
| Non-binary/Third gender or Transgender man/Transman | 1 (<1%) | 1 (<1%) | 0 (0%) |
| Race/Ethnicity |  |  |  |
| Non-Hispanic White | 720 (77%) | 447 (77%) | 273 (79%) |
| Non-Hispanic Black | 63 (7%) | 44 (8%) | 20 (6%) |
| Hispanic | 92 (10%) | 61 (10%) | 31 (9%) |
| Asian/Asian American | 26 (3%) | 9 (2%) | 17 (5%) |
| American Indian/Alaskan Native | 4 (<1%) | 4 (1%) | 0 (0%) |
| Native Hawaiian/Other Pacific Islander | 2 (<1%) | 2 (<1%) | 0 (0%) |
| Another race | 14 (2%) | 11 (2%) | 3 (1%) |
| Multiracial | 8 (1%) | 6 (1%) | 2 (1%) |
| Income |  |  |  |
| \$0 - \$49k | 206 (22%) | 117 (20%) | 89 (26%) |
| \$50K to \$99K | 362 (39%) | 232 (40%) | 130 (38%) |
| \$100K or more | 325 (35%) | 216 (37%) | 109 (32%) |
| Prefer not to say | 37 (4%) | 19 (3%) | 18 (5%) |
| Residence |  |  |  |
| Rural | 151 (16%) | 96 (16%) | 55 (16%) |
| Small city (<100,000) | 159 (17%) | 101 (17%) | 58 (17%) |
| Suburban, near a large city | 457 (49%) | 277 (47%) | 180 (52%) |
| Mid-sized city (100,000-1million) | 90 (10%) | 60 (10%) | 30 (9%) |
| large city (>1million) | 70 (8%) | 47 (8%) | 23 (7%) |
| Other | 3 (<1%) | 3 (1%) | 0 (0%) |
| Vaccination status – January 2021 |  |  |  |
| None | 765 (82%) | 463 (79%) | 302 (87%) |
| One dose | 160 (17%) | 118 (20%) | 42 (12%) |
| Two doses | 5 (1%) | 3 (1%) | 2 (1%) |
| Vaccination status – March 2021 |  |  |  |
| None (March) | 310 (33%) | 146 (25%) | 164 (47%) |
| One dose (March) | 206 (22%) | 128 (22%) | 78 (23%) |
| Two doses (March) | 414 (45%) | 310 (53%) | 104 (30%) |

**Table 2.** Bivariate correlations and odds ratios for predictor variables and COVID-19 vaccine uptake in January and March of 2021.

| | COVID-19 vaccine uptake ($\geq 1$ dose),<br>January-2021 | | | | COVID-19 vaccine uptake ($\geq 1$ dose),<br>March-2021 | | | |
| --- | --- | --- | --- | --- | --- | --- | --- | --- |
|  | *r* | *P value*<sup>*</sup> | OR | *P value* | *r* | *P value*<sup>*</sup> | OR | *P value* |
| Age | 0.16 (0.10, 0.22) | <.001 | 2.02 (1.13, 3.78) | .021 | 0.50 (0.45, 0.55) | <.001 | 10.92 (6.76, 18.05) | <.001 |
| Proportion of state with +1 dose | 0.07 (0.00, 0.13) | .239 | 1.09 (0.99, 1.18) | .063 | 0.08 (0.01, 0.15) | .070 | 0.99 (0.90, 1.10) | .909 |
| Veteran | 0.10 (0.04, 0.16) | .034 | 1.21 (0.80, 1.84) | .379 | 0.22 (0.15, 0.29) | <.001 | 1.21 (0.80, 1.84) | .366 |
| Total comorbidities | 0.09 (0.03, 0.16) | .018 | 1.10 (0.96, 1.26) | .174 | 0.17 (0.10, 0.24) | <.001 | 1.10 (0.94, 1.28) | .233 |
| Health literacy | -0.02 (-0.09, 0.04) | .948 | 1.09 (0.80, 1.44) | .557 | -0.08 (-0.15, -0.01) | .060 | 0.96 (0.72, 1.28) | .796 |
| Numeracy | 0.17 (0.10, 0.23) | <.001 | 1.48 (1.20, 1.86) | <.001 | 0.18 (0.11, 0.25) | <.001 | 0.94 (0.79, 1.13) | .509 |
| Non-Hispanic White | 0.04 (-0.02, 0.11) | .797 | 0.93 (0.58, 1.52) | .774 | 0.12 (0.05, 0.19) | <.001 | 0.98 (0.60, 1.56) | .917 |
| Worry about getting COVID-19 | 0.04 (-0.02, 0.10) | .797 | 0.84 (0.68, 1.02) | .080 | 0.12 (0.05, 0.19) | <.001 | 1.21 (0.99, 1.49) | .061 |
| COVID-19 risk perceptions | 0.11 (0.05, 0.17) | .008 | 1.35 (1.03, 1.78) | .029 | 0.13 (0.06, 0.20) | <.001 | 0.82 (0.62, 1.08) | .163 |
| Emory Vaccine Confidence | 0.15 (0.08, 0.21) | <.001 | 1.00 (0.94, 1.06) | .889 | 0.34 (0.28, 0.40) | <.001 | 1.01 (0.95, 1.07) | .726 |
| Flu vaccine important | 0.18 (0.12, 0.24) | <.001 | 1.09 (0.81, 1.51) | .581 | 0.37 (0.31, 0.43) | <.001 | 1.07 (0.83, 1.37) | .606 |
| COVID-19 vaccine important | 0.20 (0.14, 0.27) | <.001 | 1.66 (1.05, 2.66) | .033 | 0.43 (0.37, 0.49) | <.001 | 1.63 (1.15, 2.34) | .006 |
| COVID-19 vaccine intentions | 0.20 (0.14, 0.26) | <.001 | 1.29 (0.98, 1.74) | .081 | 0.42 (0.36, 0.48) | <.001 | 1.37 (1.10, 1.72) | .006 |
| Trust in healthcare | 0.08 (0.01, 0.14) | .135 | 0.89 (0.74, 1.09) | .265 | 0.25 (0.18, 0.32) | <.001 | 1.11 (0.90, 1.36) | .337 |
| (lack of) Belief in science | -0.15 (-0.21, -0.08) | <.001 | 0.87 (0.74, 1.02) | .089 | -0.25 (-0.31, -0.18) | <.001 | 0.84 (0.72, 0.99) | .041 |
| Belief in conspiracy theories | -0.11 (-0.17, -0.05) | .008 | 1.10 (0.81, 1.47) | .532 | -0.25 (-0.32, -0.19) | <.001 | 1.03 (0.80, 1.33) | .823 |
| Conservative beliefs | -0.02 (-0.09, 0.04) | .948 | 1.04 (0.92, 1.19) | .500 | -0.02 (-0.09, 0.05) | .610 | 1.05 (0.91, 1.20) | .514 |
| Maximizing | 0.05 (-0.01, 0.12) | .473 | 0.97 (0.85, 1.11) | .682 | 0.14 (0.07, 0.21) | <.001 | 1.11 (0.97, 1.27) | .128 |

*r* represents the bivariate correlation coefficient, OR represents the odds ratio.
95% confidence intervals (CIs) are shown in parentheses.
\*Holm-Bonferroni correction applied.

**Table 3.** Reasons for not getting a COVID-19 vaccine.

|  | *Unvaccinated respondents who were*<br>*not interested in getting a COVID-19 vaccine* |  |  | *All respondents from wave 1 and all*<br>*unvaccinated respondents in waves 2 and 3* |  |  |
| --- | --- | --- | --- | --- | --- | --- |
|  | December, 2020<br>(N=69) | January, 2021<br>(N=69) | March, 2021<br>(N=69) | December, 2020<br>(N=2085) | January, 2021<br>(N=1043) | March, 2021<br>(N=361) |
| Safety – no. (%) |  |  |  |  |  |  |
| Concerns about side effects | 22 (32) | 14 (20) | 20 (29) | 686 (33) | 159 (15) | 65 (18) |
| Concerns about vaccine development process | 5 (7) | 16 (23) | 13 (19) | 235 (11) | 76 (7) | 27 (8) |
| Worried about getting COVID-19 from the vaccines | 3 (4) | 2 (3) | 2 (3) | 74 (5) | 16 (2) | 7 (2) |
| I don't like needles | 4 (6) | 1 (1) | 2 (3) | 83 (4) | 12 (1) | 11 (3) |
| Efficacy – no. (%) |  |  |  |  |  |  |
| Doubt vaccine efficacy | 4 (6) | 7 (10) | 4 (6) | 98 (5) | 19 (2) | 7 (2) |
| Low COVID-19 threat – no. (%) |  |  |  |  |  |  |
| I won't get COVID-19 even if I don't get the vaccine | 4 (6) | 5 (7) | 5 (7) | 63 (3) | 12 (1) | 14 (4) |
| COVID-19 is not as serious as some people say | 6 (9) | 3 (4) | 6 (9) | 40 (2) | 12 (1) | 11 (3) |
| I do not think I'll get very sick if I get COVID-19 | 1 (1) | 3 (4) | 1 (1) | 55 (3) | 8 (.8) | 8 (2) |
| Personal beliefs – no. (%) |  |  |  |  |  |  |
| Against religious/philosophical beliefs | 6 (9) | 3 (4) | 4 (6) | 34 (2) | 7 (.7) | 10 (3) |
| Distrust of big Pharma/government | 5 (7) | 5 (7) | 4 (6) | 97 (5) | 21 (2) | 16 (4) |
| Other reasons – no. (%) |  |  |  |  |  |  |
| Already had COVID-19 | 1 (1) | 3 (4) | 2 (3) | 28 (1) | 15 (1) | 5 (1) |
| No specific reason/multiple reasons/Unsure | 1 (1) | - | 3 (4) | 13 (.6) | 7 (.7) | 5 (1) |
| Did not respond | 5 (7) | 5 (7) | - | 185 (9) | 69 (7) | 1 (.3) |
| I plan to get the vaccine | - | 1 (1) | 1 (1) | 309 (15) | 599 (57) | 166 (46) |
| Access/Availability/Cost | - | 1 (1) | - | 36 (2) | 5 (.5) | 2 (.6) |
| Medical reasons (e.g., allergies) | 2 (3) | - | 2 (3) | 49 (2) | 6 (.6) | 6 (2) |

### Procedure and measures

Over a four-month period (December-2020 to March-2021), respondents completed three surveys (available in the appendix) which consisted of questions about their current behaviors, well-being, healthcare experiences, and attitudes regarding the COVID-19 pandemic. Both the January and March surveys also contained short message-based experiments regarding the COVID-19 vaccines which have been published elsewhere.^21,22^

Descriptions of all the measures included in the analyses are available at: https://rpubs.com/AThorpe/CV19VaxUptakeMeasures.

#### Primary outcome measure

Self-reported vaccination status in January and March-2021, measured using a single question with three options (0=No; 1=Yes, 1 dose; 2=Yes, 2 doses). As responses 1 and 2 indicated receiving at least one dose of a COVID-19 vaccine, they were considered vaccinated for analyses (0=Not vaccinated; 1=Vaccinated).

#### Early vaccine eligibility

Respondents’ age and the total number of comorbidities^23^ were included based on recommendation by the Centers for Disease Control and Prevention (CDC) for these populations to be offered vaccines first.^24^ As the speed of vaccine distribution within each state may affect vaccine availability for those eligible we also included the proportion of each state that had received at least one dose of a COVID-19 vaccine (retrieved from publicly available data: https://www.kff.org/coronavirus-covid-19/issue-brief/state-covid-19-data-and-policy-actions/). Veteran status (0=non-Veteran; 1=Veteran) was also included given the involvement of the U.S. Department of Veterans Affairs in the distribution of COVID-19 vaccines following their authorization.^25^

#### Demographic factors

We included respondents health literacy,^26^ numeracy^27,28^ and Race/Ethnicity (dummy coded as 0= any other race/ethnicity; 1= non-Hispanic white).

#### Psychological factors

We included respondents’ worries and risk perceptions about COVID-19, the Emory Vaccine Confidence Index,^29^ perceived importance of Flu and COVD-19 vaccines, COVID-19 vaccine intentions, trust in healthcare,^30^ (lack of) belief in science,^31^ belief in conspiracy theories,^32^ political views, and the single-item maximizer-minimizer elicitation question (the MM1; which measures preference for either waiting or taking action in medical situations where it is unclear whether action is needed).^33^

### Analysis

All the analyses were conducted in R Studio Version 1.4.1106.^34^ We used the “psych” package^35^ to run bivariate correlations between our predictor variables and vaccine uptake. Using the “stats” package,^36^ we ran a multiple logistic regression model to test whether the early vaccine eligibility and demographic factors predict getting at least one dose of a COVID-19 vaccination in January-2021. Using a hierarchical approach, we then included the psychological factors to the original model. We then repeated this analytical approach with receiving at least one dose of a COVID-19 vaccine in March-2021 as the dependent variable.

## Results

In the regression models which only included the early vaccine eligibility and demographic factors, we found that older age (OR*=*2.54[95%CI=1.47 – 4.65], *p*=.001), the proportion of the state vaccinated (1.09[1.00–1.19], *p=*.041), increased number of comorbidities (OR=1.17[1.03– 1.33], *p*=.034), and higher numeracy (1.59[1.30–1.97], *p*<.001) predicted vaccine uptake in January. Older age (9.10[6.01–14.03], *p*<.001), increased number of comorbidities (1.19[1.04– 1.35], *p*=.011), and higher numeracy (1.18[1.01–1.37], *p*=.035) were significant predictors of later vaccine uptake (in March).

After including the psychological variables, older age remained a predictor of vaccine uptake in both January and March (Figure 1). Higher numeracy, higher COVID-19 risk perceptions, and believing that it is important for all adults to get the COVID-19 vaccine were also predictors of vaccine uptake in January. In March, believing that it is important for all adults to get the COVID-19 vaccine, prior intentions to get a COVID-19 vaccine, and general belief in science predicted vaccine uptake alongside older age.

**Figure 1.**
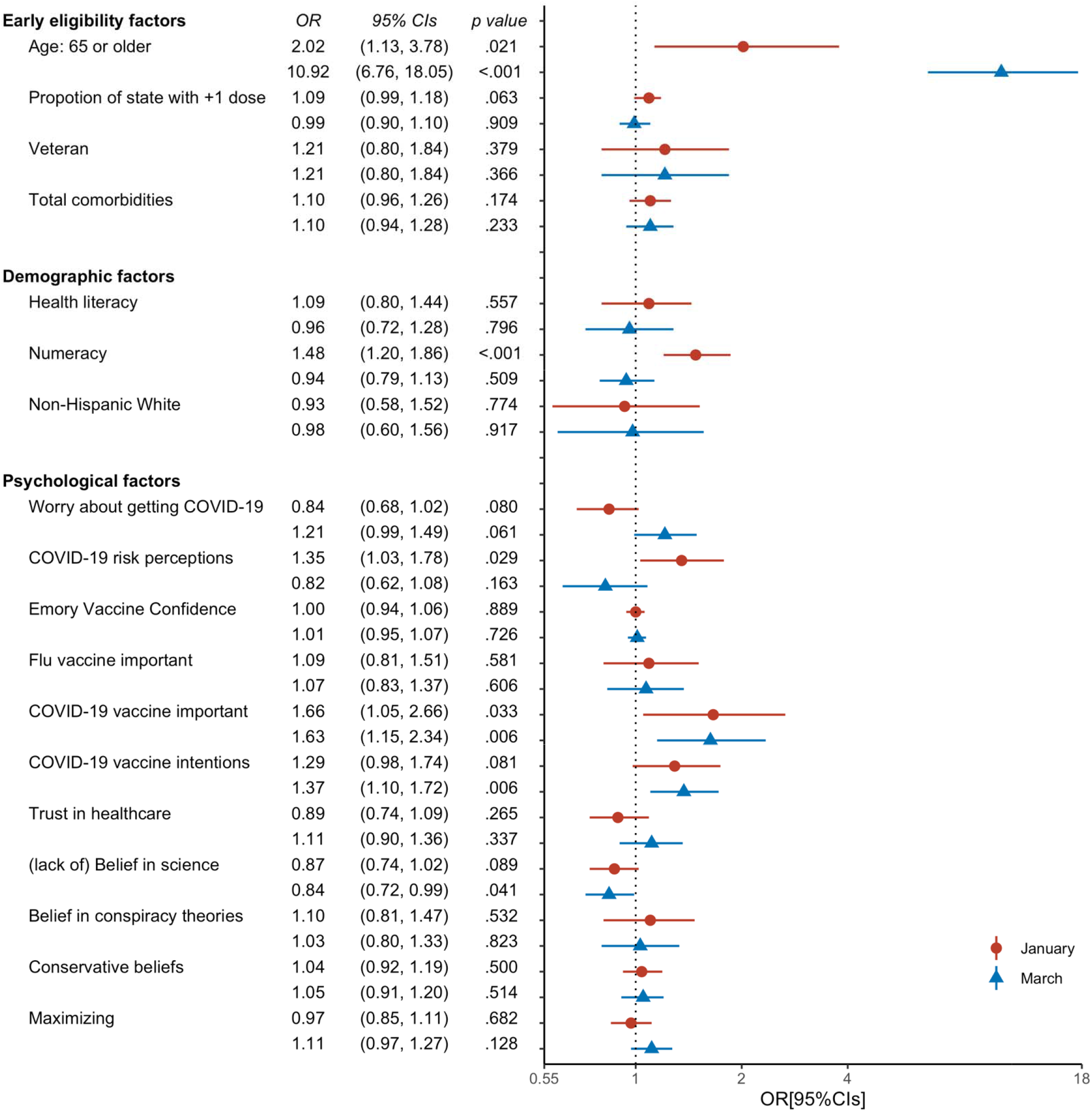
Odds ratios with 95% confidence intervals for predictors of respondents’ vaccination status in both January and March 2021. Reference categories were 64 or younger (for Age), non-Veteran (for Veteran), and any other Race/Ethnicity (for Non-Hispanic White).

In the March-2021 survey, a total of 310 respondents (33% of the total sample) had not received a vaccine. Almost a quarter of those respondents (69 (22%)) reported that they did not want to get one. Among the 69 unvaccinated respondents who did not want to receive a COVID-19 vaccine, concerns about possible side effects and the vaccine development process were the most frequently endorsed reason for not getting vaccinated (Table 1). Other reasons for not getting vaccinated included not believing COVID-19 poses a serious threat, personal beliefs (e.g., religious and philosophical) that conflicted with getting vaccinated, and distrust of the institutions involved with promoting vaccines (e.g., pharmaceutical companies and the government). A few respondents cited doubts about the efficacy of the vaccines and a very small proportion reported access issues (e.g., not having enough time or vaccines being unavailable) as reasons for not getting vaccinated.

In a further exploratory analyses, we found that younger age (0.27[0.09–0.77], *p*=.016), believing it is not important for all adults to get a COVID-19 vaccine (0.15[0.08–0.26], *p*<.001), low trust in healthcare (0.59[0.36–0.95], *p*=.032), and preferring to watch and wait in medical situations where it is not clear whether or not medical action is necessary (0.66[0.48–0.89], *p*=.007), were significant predictors of being unvaccinated and not wanting to receive a COVID-19 vaccine by March-2021 (Figure 2).

**Figure 2.**
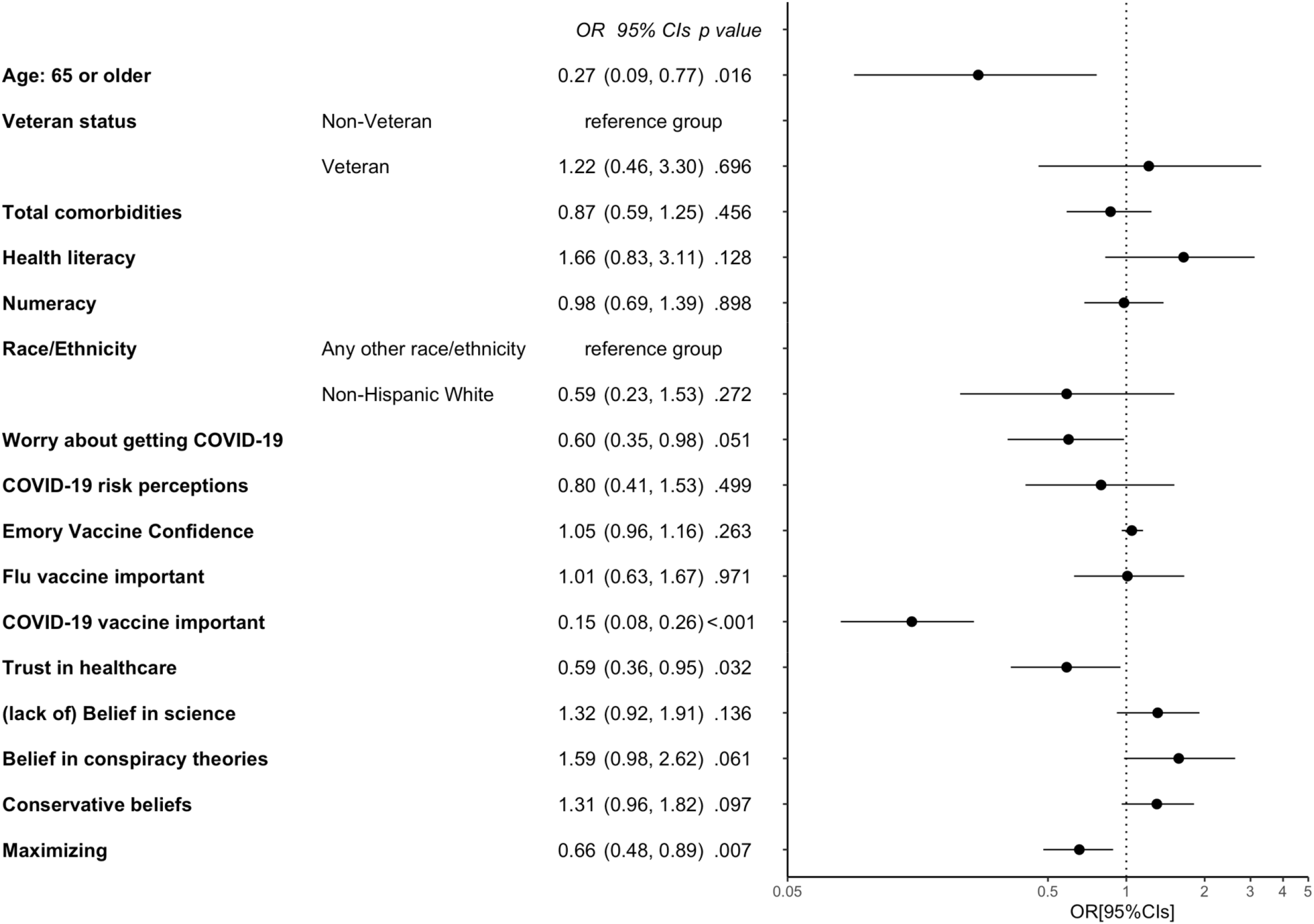
Odds ratios with 95% confidence intervals for predictors of being unvaccinated and not wanting a COVID-19 vaccine by March 2021.

## Discussion

The aim of the present, longitudinal study was to identify key predictors of, and objections to, COVID-19 vaccine uptake. We found that older age, higher numeracy, higher COVID-19 risk perceptions, and positive attitudes towards COVID-19 vaccines were important predictors of early vaccine uptake (by January-2021). As the rollout progressed, the influence of numeracy and risk perceptions remitted and we found that only older age, positive attitudes towards COVID-19 vaccine, and intentions to receive a COVID-19 vaccine were significant predictors of later vaccine uptake (by March-2021).

Older age was the strongest predictor of vaccine uptake for both timepoints, which reflects its emphasis as key criterion for early vaccine eligibility by the CDC.^24^ The combination of numeracy, risk perceptions, and attitudes towards COVID-19 vaccines as predictors of early vaccine uptake supports prior research demonstrating that assessment of the risks and benefits offered by vaccination as well as the threat of the disease that the vaccine protects against have a substantial influence on whether or not someone is likely to get vaccinated.^37,38^ While clear communication about the risks and benefits associated with the vaccine and the threat posed by the disease is crucial at all times, these findings suggest that it may be particularly effective at encouraging uptake during the early stages of rollouts and for novel vaccines and diseases.

Our findings offer important evidence that attitudes and intentions towards COVID-19 predict uptake and provide validation for the many studies that have used these measures as a proxy for vaccination uptake.^2–4,6–9,11,12^ In fact, of our respondents who were 65 years or older, only 8% of those who reported that they intended to vaccinate had not done so by the March-2021 survey. We did not consider respondents younger than 65 years old as recommendations to make COVID-19 vaccines available to all adults aged between 16 and 65 were only announced in March 17, 2021 so it was unlikely they would have had the opportunity to be vaccinated.^39^ Taken together, these findings reinforce the need to develop effective strategies for addressing people’s concerns and negative attitudes towards COVID-19 vaccines in order to increase uptake.

The findings from the present study may also contribute to informing health communication efforts aimed at those least likely to get a COVID-19 vaccine. Around 10% of the respondents in our study had not been vaccinated at the time of the final survey in March and indicated that they did not intend to do so. These respondents tended to be younger, had negative views about the COVID-19 vaccines for adults, low trust in healthcare, and preferred to watch and wait before taking action in medical situations where there is clinical equipoise on whether action is necessary. In addition, the most important reasons given by these respondents for not getting a COVID-19 vaccine focused on safety concerns (particularly regarding side effects and the development process), beliefs that COVID-19 is not a serious threat, personal beliefs conflicting with vaccination and distrust of institutions involved with the vaccines. Our findings are aligned with prior studies on the reasons given by people who are hesitant towards or refuse COVID-19 vaccines,^3,40–42^ and offer an empirical basis for targeting public health messages to those who are least likely to vaccinate and tailoring messages to address their concerns. As these beliefs are often deeply held and traditional models of health communication have been largely ineffective at addressing them,^21,43^ we encourage health researchers and communicators to move beyond such traditional models of information provision and instead generate alternative strategies for addressing the concerns of those who are reluctant to get vaccinated.

One limitation of the study is that the findings rely on the accuracy and consistency of respondents’ self-reported data over the duration of the survey period. Although self-reports have been shown to be highly concordant with healthcare utilization and vaccine records,^44,45^ replication of these findings with a method for confirming respondents’ reported vaccine uptake would increase confidence in these findings.

Furthermore, our sample consisted of Veteran and non-Veteran respondents who were unique in being sufficiently motivated and able to complete three online surveys during the pandemic and therefore are not representative of the general U.S. population. The finding that Veteran status did not predict vaccine uptake at either time point was surprising given the efforts and widespread outreach of the U.S. Department of Veterans Affairs in supporting COVID-19 vaccine distribution.^46^ However, it is likely that the greater proportion of older adults in the Veteran sample compared to the non-Veteran sample may have limited our ability to observe a significant effect of Veteran status in the full model. In addition, our sample was overrepresented by respondents without many health conditions (70% reported ≤ 1 health condition), with high health literacy (94% of respondents reported high health literacy), and who identified as non-Hispanic White (78%). The unique makeup of our sample may also explain why older age and numeracy were the only early eligibility and demographic factors associated with vaccine uptake.

Despite these limitations, the findings from the present study offer important insights regarding the predictors of vaccine uptake during the early stages of the COVID-19 vaccine rollout in the US, which can help guide health communications and public outreach. Our findings reinforce the need for developing effective strategies for promoting positive attitudes and intentions towards vaccines to promote uptake. A major strength of our study is that we were able to cover the initial stages of the COVID-19 vaccine distribution. However, given the changes observed between January and March and the unique characteristics of our sample, further studies are needed to re-evaluate the key predictors of vaccine uptake as the rollout progresses and with wider representation.

## Data Availability

Data and study materials are available from the first author upon request.

## Acknowledgements

The pre-registration document associated with this manuscript is available at: https://aspredicted.org/MKS_HRZ. The views expressed in this paper are those of the authors and do not necessarily represent the position or policy of the U.S. Department of Veterans Affairs or the United States Government.

## Footnotes

a In our pre-registration (https://aspredicted.org/MKS_HRZ) we erroneously stated that we would expect “lower health literacy, lower numeracy” to be associated with vaccine uptake. This was an error and is therefore corrected in the manuscript.

